# Subthreshold intracortical microstimulation of human somatosensory cortex enhances tactile sensitivity

**DOI:** 10.1101/2024.06.21.24309202

**Authors:** Luke E. Osborn, Breanne Christie, David P. McMullen, Victoria Arriola, Tessy M. Thomas, Ambarish S. Pawar, Robert W. Nickl, Manuel A. Anaya, Brock A. Wester, Charles M. Greenspon, Gabriela L. Cantarero, Pablo A. Celnik, Sliman J. Bensmaia, Jeffrey M. Yau, Matthew S. Fifer, Francesco V. Tenore

## Abstract

Intracortical microstimulation (ICMS) of the somatosensory cortex activates neurons around the stimulating electrodes and can elicit tactile sensations. However, it is not clear how the direct activation of cortical neurons influences their ability to process additional tactile inputs originating from the skin. In a human implanted with chronic microelectrode arrays in both left and right somatosensory cortices, we presented mechanical vibration to the skin while simultaneously delivering ICMS and quantified the effects of combined mechanical and electrical stimulation on tactile perception. We found that subthreshold ICMS enhanced sensitivity to touch on the skin, as evidenced by a reduction in vibrotactile detection thresholds (median: −1.5 dB), but subthreshold vibration did not systematically impact the detectability of ICMS. Suprathreshold vibration led to an increase in ICMS thresholds (median: 2.4 dB) but suprathreshold ICMS had little impact on vibrotactile thresholds. The ICMS-induced enhancement of vibrotactile sensitivity was location dependent with the effect size decreasing as the projected field of the stimulating electrode and the locus of vibratory stimulation became farther apart. These results demonstrate that targeted microstimulation of cortex alone can focally enhance tactile sensitivity, potentially enabling restoration or strengthening of retained tactile sensations after injury.

## INTRODUCTION

Our interactions with objects rely heavily on the sense of touch, which can be compromised for individuals living with sensorimotor deficits, such as those resulting from spinal cord injury, stroke, or amputation. There is evidence that introducing low levels of mechanical (*1*, *2*) or electrical (*3*) noise at the peripheral level can boost the detectability of a haptic signal on the skin or even provide functional benefit such as improving balance (*4*), all of which are potentially explained by stochastic resonance within somatosensory networks. Stochastic resonance, wherein a subthreshold signal is made more detectable by the introduction of noise in that signal, has been repeatedly demonstrated in biological sensory systems (*5*, *6*) and explored using theoretical frameworks (*7*).

Subthreshold skin vibration has been reported to improve touch sensitivity (*8*) and motor function (*9*) in stroke survivors. The effect of enhanced sensory detection during the presence of low amplitude noise is also apparent in the auditory system using either acoustic or electrical stimulation of the auditory nerve or brainstem (*10*), although not all reports have found consistent effects (*11*). Similarly, direct activation of neurons through low-amplitude intracortical microstimulation (ICMS) has been shown to improve detection of visual stimuli (*12*) and ICMS-evoked artificial tactile percepts (*13*) in non-human primates.

While ICMS of somatosensory cortex is often used to elicit tactile sensations in humans (*14–16*), a large portion of the target population for implanted microelectrode arrays retains some degree of tactile sensation – including individuals with incomplete spinal cord injury or motor paralysis. Therefore, it is important to understand the effects of artificial electrical activation of cortical neurons on the ability to perceive and process natural tactile inputs originating from the skin. While it is possible to simultaneously perceive and differentiate ICMS-evoked sensations from naturally occurring tactile inputs at the same location of the hand (*17*), it is unknown how ICMS of the somatosensory cortex interacts with existing touch pathways and perception.

To fill this gap, we investigated the ability of a human participant with microelectrode arrays chronically implanted in putative Brodmann area 1of the somatosensory cortex to detect skin vibrations or ICMS alone or in the presence of one another. To determine if the interactions depended on stimulation location, the ICMS and vibratory stimuli were designed to activate either overlapping or spatially distinct regions of the hand. This approach was unique because we delivered somatosensory information to two different targets in the somatosensory pathway (i.e., cortex and skin), yet both inputs could be perceived simultaneously as tactile sensation on the hand. Simultaneous ICMS and skin vibration activate overlapping neural populations in the somatosensory cortex (*18*), and therefore provided an opportunity for the two stimulation signals to interact given the placement of implanted electrodes in the human participant (*16*, *19*).

We first found that subthreshold ICMS delivered to somatosensory cortex resulted in enhanced vibration detection sensitivity. Second, the degree of the enhanced sensitivity depended on the degree of overlap of the electrically- and mechanically-activated regions of the hand. Third, our results revealed that subthreshold vibration did not systematically impact the detectability of ICMS. Fourth, suprathreshold vibrations reduced sensitivity to ICMS while the converse was not true (that is, suprathreshold ICMS did not have a consistent effect on vibrotactile sensitivity). These results demonstrate that artificial touch elicited by electrical activation of somatosensory cortex interacts in complex ways with its mechanical counterpart, and that direct cortical stimulation can be used to alter perceptual sensitivity of tactile stimulation on the skin.

## RESULTS

Microelectrode arrays were chronically implanted in the motor and somatosensory cortices in both hemispheres of a person with a spinal cord injury. As previously reported, we found that ICMS delivered to the somatosensory cortex elicited artificial tactile sensations in the participant’s hands (**Fig. 1A-B**) (*16*), often described as pressure experienced on a patch of skin. In addition, the location of the evoked sensation depended on the electrode through which stimulation was delivered, following the progression of the somatosensory homunculus, as has been previously shown (*16*, *20*). The participant retained some tactile sensitivity in his hands and in trials that involved cutaneous vibration, he described sensations evoked by vibratory stimulation as “buzzing” or “tingling.”

**Fig. 1:**
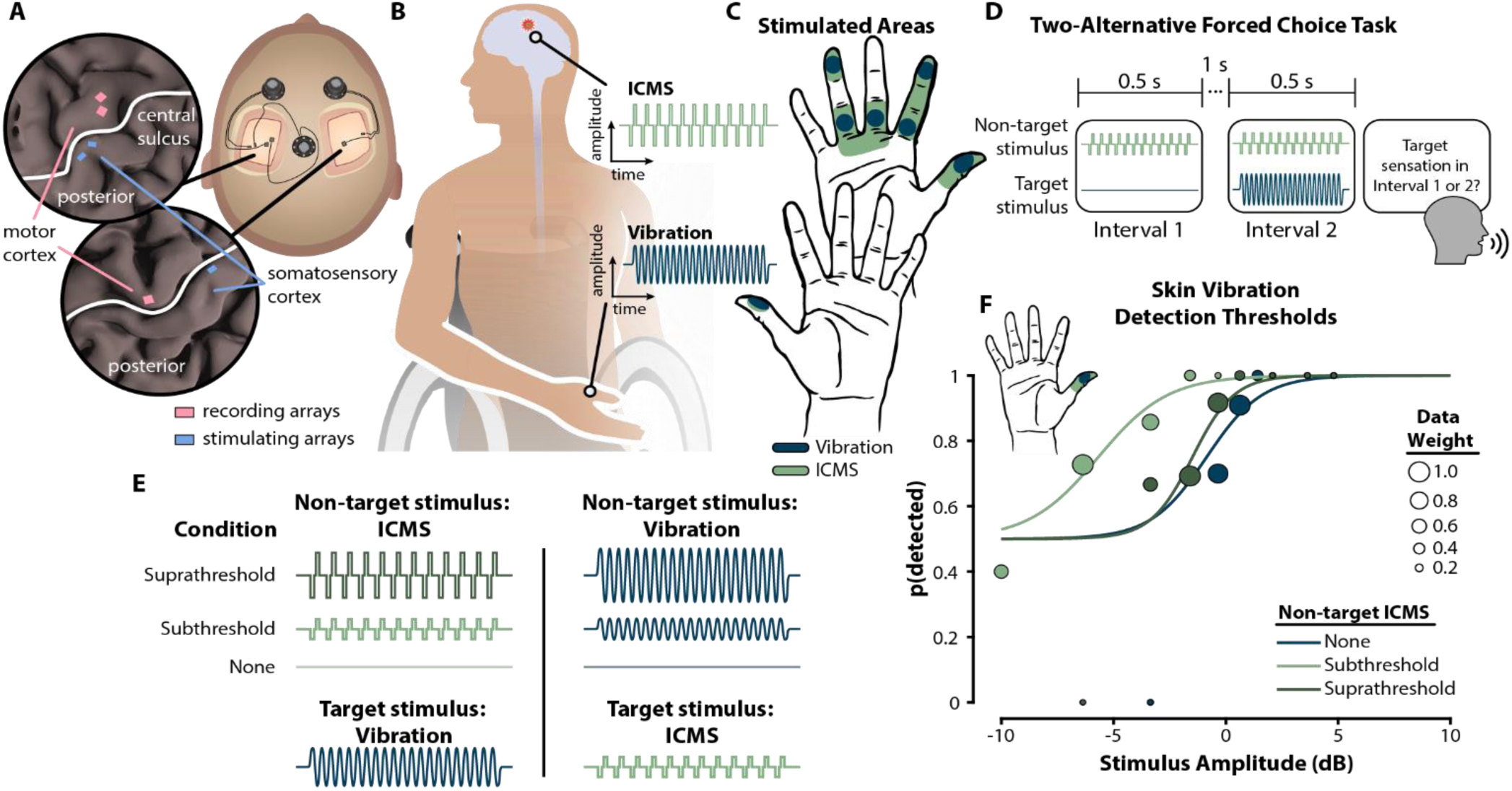
Brain stimulation modulates tactile sensitivity. **(A)** Chronic microelectrode arrays implanted in motor and somatosensory cortices of both hemispheres covered hand regions. **(B)** Intracortical microstimulation (ICMS) of the somatosensory cortex and vibratory stimuli on the skin produced tactile sensations in the hands of a human participant. **(C)** The detection thresholds for vibratory stimuli and ICMS-elicited touch were tested in multiple locations across both hands. **(D)** In a two-alternative forced choice task to measure detection thresholds, the target stimulus was randomly delivered in one of two trial intervals and the participant reported whether they detected the target in the first or second interval. A non-target stimulus was delivered in both intervals. **(E)** We tested multiple conditions where the non-target stimulus was either suprathreshold or subthreshold. The non-target modality was always different than the target modality stimulus. **(F)** On the right thumb fingertip, delivery of a subthreshold ICMS reduced the detection threshold for skin vibration by 5.1 dB. Marker size indicates the relative weights, based on number of presentations, of each stimulus level used for fitting the psychometric curve.

Detection thresholds were measured using a two-interval, two-alternative forced choice (2AFC) adaptive staircase method. Vibratory thresholds were measured at multiple sites across both hands (**Fig. 1C**; *n* = 7). ICMS thresholds were measured on electrodes with projection fields that overlapped with the vibratory threshold measurement sites (**Fig. 1C**). Vibratory and ICMS thresholds were measured in the absence and presence of a “non-target” stimulus, which was delivered in the other modality simultaneously with the target stimulus (**Fig. 1D-E**). The non-target stimulus was either subthreshold (imperceivable) or suprathreshold (perceivable) (**Fig. 1E**). Unless stated otherwise, the site of vibratory stimulation coincided with the projected field of ICMS.

### Subthreshold cortical stimulation increases vibrotactile sensitivity

We related the amplitude of the target stimulus to its detectability and then evaluated detectability with and without the non-target stimulus. We found that subthreshold ICMS increased the sensitivity to vibration compared to the baseline condition without ICMS, as indicated by a leftward shift in the psychometric function (**Fig. 1F**). Across sites on both hands and across visits, we found that subthreshold ICMS systematically and significantly increased vibrotactile sensitivity (median: −1.5 dB, *p* = 0.002) (**Fig. 2**). A bootstrap analysis revealed that out of 17 experiment sessions with the subthreshold ICMS condition, 12 (70.6%) of them showed a significant shift in vibratory detection threshold from baseline (**Fig. S1**). In contrast, delivery of subthreshold vibration minimally impacted ICMS detection (median threshold shift: 0.2 dB, *p* = 0.36) with 3/10 (30%) of the experimental values being significantly different compared to baseline levels (**Fig. 2B**). Statistical significance of each data point was determined by the proportion of simulated detection thresholds that were above or below the baseline value (see Materials and Methods).

**Fig. 2:**
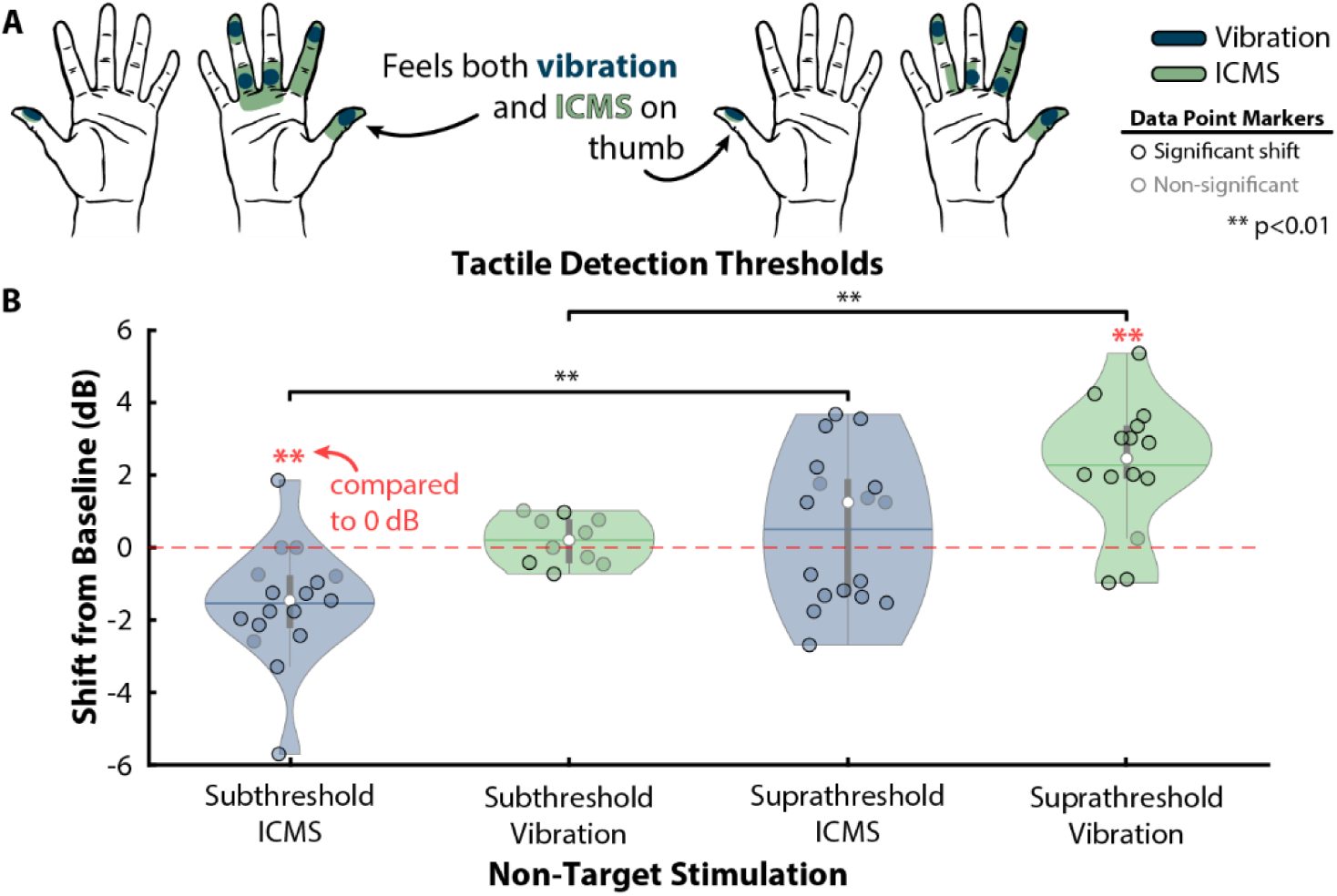
Subthreshold cortical stimulation enhances tactile sensitivity. **(A)** Skin vibration was applied to six different parts of the hand during simultaneous delivery of either a subthreshold or a suprathreshold non-target ICMS signal to neural tissue assumed to encode the same hand region. During suprathreshold conditions, the participant perceived tactile sensations on the target hand sites from both the vibration and ICMS. **(B)** Subthreshold ICMS led to a significant reduction (median: −1.5 dB, *p* = 0.002) in vibration detection threshold, and this increased sensitivity was consistent across all hand regions. Providing suprathreshold ICMS did not significantly alter vibrotactile thresholds with respect to the baseline value (*p* = 0.27). Subthreshold non-target vibration did not alter ICMS detection thresholds (median: 0.2 dB, *p* = 0.36); however, ICMS thresholds did significantly increase during simultaneous delivery of suprathreshold vibration signals (median: 2.4 dB, *p* = 0.001). The violin plot whiskers represent the minimal and maximal values; the vertical lines indicate the first and third quartiles; the horizontal lines are means; the white dots are the medians.

When considering only the magnitude of the shift in detection thresholds, both subthreshold ICMS and vibration resulted in significant deviations from baseline (*p* < 0.01, **Fig. S2**) indicating that either non-target stimuli modality impacts detectability of the target stimulation; however, the effects of subthreshold skin vibrations on ICMS detection are notably less consistent.

To determine if the projected field size or ICMS amplitude played a role in the observed shift in vibrotactile sensitivity during subthreshold ICMS, we calculated the Pearson correlation coefficient to the shift in detection thresholds. There was no significant correlation between the projected field size or the amplitude of ICMS and the shift in vibration detection thresholds (**Fig. S3**).

### Suprathreshold skin vibration decreases ICMS sensitivity

Next, we examined the effects of suprathreshold ICMS on vibrotactile sensitivity and those of suprathreshold vibration on ICMS sensitivity. We found that suprathreshold ICMS did not have a consistent effect on vibrotactile sensitivity (*p* = 0.27): suprathreshold ICMS elevated vibrotactile thresholds during some sessions and reduced vibrotactile thresholds in other sessions (**Fig. 2A**). Indeed, suprathreshold ICMS was associated with significant threshold shifts in 14/17 sessions (82.4%), an occurrence greater than expected by chance (binomial test *p* < 0.001). Accordingly, considering only the magnitude and not the direction of shift from baseline detection thresholds, there was a significant effect (*p* < 0.001) with suprathreshold ICMS during skin vibration detection tasks (**Fig. S2**).

In contrast, suprathreshold vibration consistently reduced the participant’s sensitivity to ICMS, as evidenced by consistent increases in threshold (median shift: 2.4 dB, *p* = 0.001, **Fig. 2B**). Of the experimentally measured detection thresholds, 12/14 (85.7%) were significantly different than baseline levels. To determine if the amplitude of the suprathreshold non-target stimulus affected detection thresholds of the target stimulus, we performed experimental sessions with low, high, or random non-target stimulus amplitudes but found statistically indistinguishable effects on detectability (*p* > 0.05, **Fig. S4**). We also measured the magnitude of the shift in detection thresholds from baseline to determine if the size of the effect was dependent on the amplitude of suprathreshold non-target stimuli. For both suprathreshold ICMS and vibration conditions, there was no significant difference between various non-target stimulation amplitudes (*p* > 0.05, **Fig. S5**).

To determine the relationship between subthreshold and suprathreshold non-target stimulation, we calculated the Pearson correlation coefficient (*r*) between the shift in detection thresholds for each condition. We found a slightly positive, yet statistically significant (*p* < 0.05), correlation between the two non-target stimulation conditions for both vibration and ICMS detection tasks (**Fig. S6A-B**). The correlation between the two conditions did not persist when considering only the magnitude of shifts in vibration sensitivity during non-target ICMS; however, there was still a correlation for the ICMS detection task (**Fig. S6C-D**).

### Proximity of target and non-target stimuli modulates degree of enhanced tactile sensitivity

In the above analyses, we restricted comparisons to conditions in which ICMS was delivered through an electrode whose projected field overlapped with the location at which the vibratory stimulus was delivered. Under these conditions, we reasoned that vibration and ICMS activated overlapping populations of neurons given the overlap of receptive and projected fields (*20*).

We next asked whether the enhanced vibrotactile sensitivity observed in the presence of a subthreshold ICMS distractor was predicated upon the overlap of the projected field of ICMS-evoked sensation and the vibration location. To this end, we delivered ICMS to a region of somatosensory cortex whose projected field *did not* overlap with the locus of vibratory stimulation and compared the vibratory detection thresholds to when subthreshold ICMS was delivered to a somatotopically matched region of somatosensory cortex (**Fig. 3A**).

**Fig. 3:**
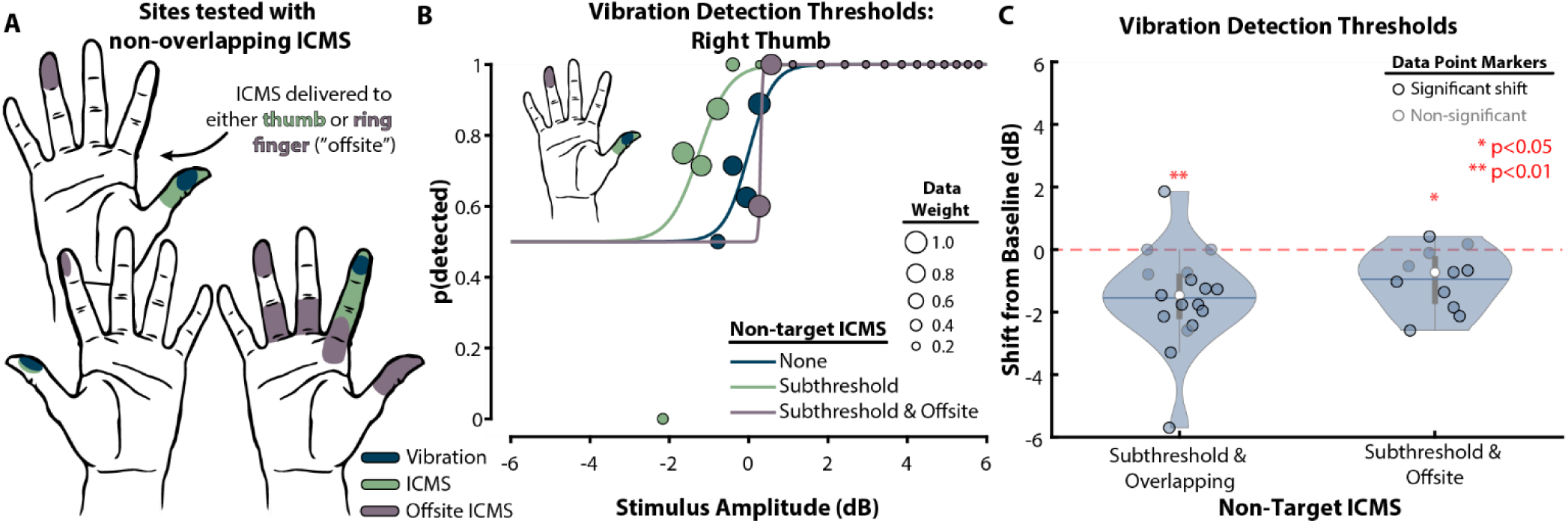
Enhanced tactile sensitivity depends on relative location of subthreshold ICMS. **(A)** We delivered ICMS to neural regions that represented hand areas that did not overlap with the sites of vibratory stimuli. When vibrating the thumb on either the right or left hand, we simultaneously provided non-overlapping ICMS to the ring finger and left index finger, respectively. During vibration of the right index finger, we delivered ICMS to non-overlapping regions ranging from near to far from the target location (i.e., index fingertip). **(B)** When subthreshold ICMS to the ring finger acted as the non-target stimulus, the vibration detection threshold on the right thumb (i.e., an “offsite” region of the hand) was not altered; however, subthreshold ICMS delivered to the right thumb led to a decrease in the vibration detection threshold. **(C)** Subthreshold ICMS delivered to non-overlapping (“offsite”) regions of the hand led to a slight decrease in vibration detection thresholds (median: −0.7 dB, *p* < 0.05) although this decrease in threshold was stronger when subthreshold ICMS was delivered to an overlapping region (median: −1.5 dB, *p* < 0.01), suggesting that relative location of ICMS is important for enhancing tactile sensitivity.

We found that the size of subthreshold ICMS effects depended on the somatotopic overlap of ICMS and vibrotactile stimuli (**Fig. 3B-C**). The effect on detection thresholds was smaller (robust Cohen’s effect size *d_r_* = −0.72) when subthreshold ICMS was delivered to non-overlapping regions of the hand compared to when there was somatotopic overlap (*d_r_ =* −1.28). However, across all sites, non-overlapping ICMS produced a significant decrease in vibrotactile thresholds (median: −0.7 dB, *p* < 0.05; 8/11 (72.7%) significantly different than baseline). Non-overlapping ICMS effects were nominally weaker than in the overlapping ICMS effects (median: −1.5 dB), but these differences did not achieve statistical significance (**Fig. 3C***; p* = 0.20, *d_r_* = 0.48).

Next, we further assessed the dependence of the enhancement of vibrotactile sensitivity on somatotopic alignment, which presumably is related to neural activation overlap. To do this, we analyzed changes in tactile detection sensitivity as a function of the physical cortical distance between the electrode delivering ICMS and the location of the electrode that, when stimulated, evoked a projected field that maximally overlapped with the vibration stimulus (**Fig. 4A**).

**Fig. 4:**
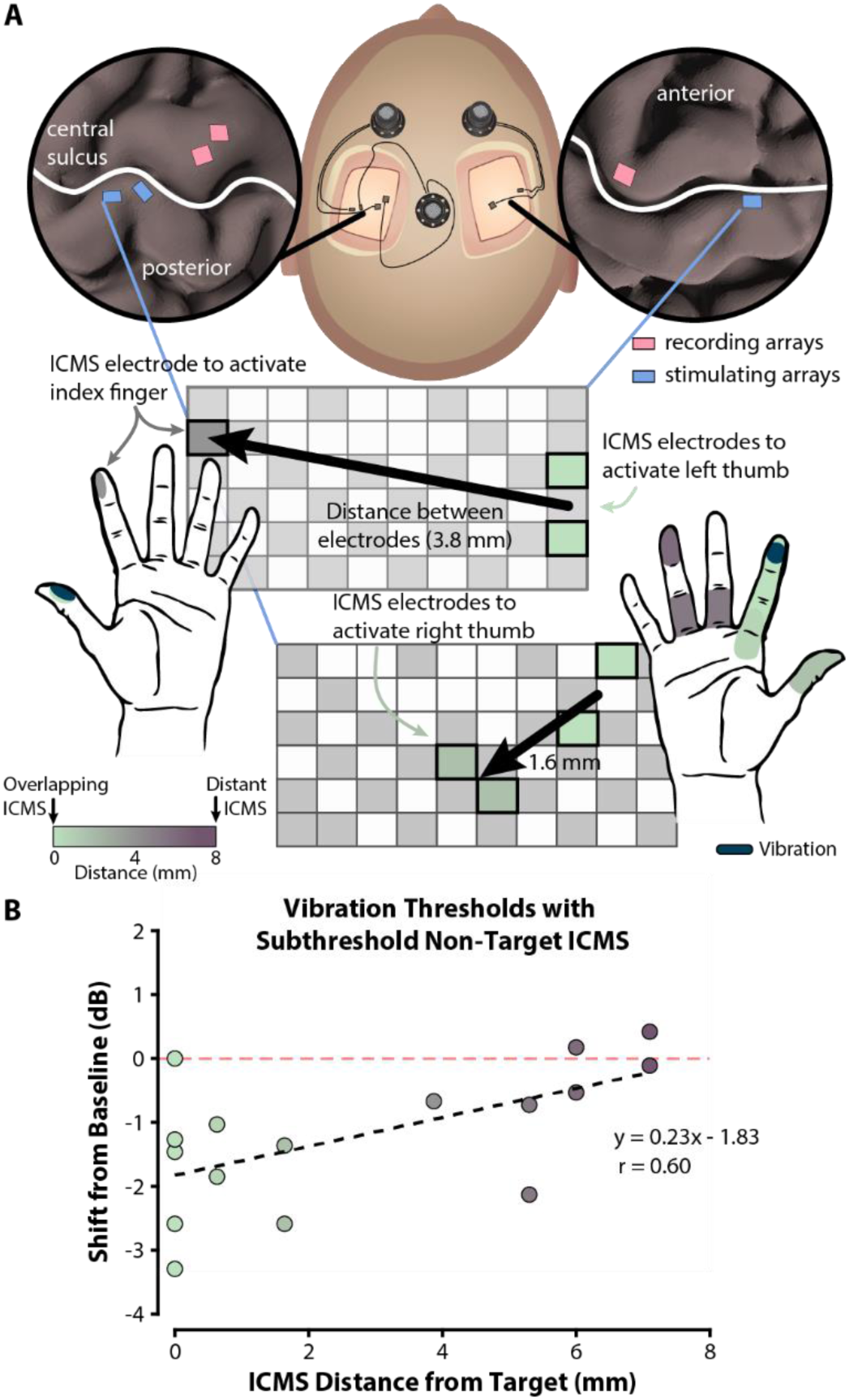
Enhanced tactile sensitivity decreases with cortical distance between stimuli. **(A)** Linear distances between stimulation sites were approximated based on the centroid location of each electrode group on the microelectrode array. Overlapping ICMS (i.e., delivered to a site that somatotopically aligned with skin vibration) was defined as being 0 mm from the target. A range of sites were tested to evaluate the effect of subthreshold ICMS location on vibration sensitivity. **(B)** Vibration detection thresholds were maximally reduced when subthreshold ICMS was applied to a neural region representing the same region of the hand. The enhanced sensitivity effect degraded as ICMS moved away from the target neural region at a rate of 0.23 dB / mm (*p* < 0.05). There was a correlation between reduced vibration thresholds and ICMS distance from the target (*r* = 0.6, *p* < 0.05).

We found that as the distance between the electrode delivering ICMS and the electrode overlapping with the vibratory stimulus increased, the degree of sensitization decreased (**Fig. 4B**), yielding a significant correlation between distance and threshold shift (*p* < 0.05). We estimate that the effect of ICMS on vibratory sensitivity vanishes for distances greater than ~7 mm (**Fig. 4B**).

### Baseline detection thresholds do not shift within a visit

Having observed that ICMS can influence sensitivity to vibration stimuli and vice versa, we assessed whether these effects might reflect shifts in sensitivity caused by perceptual adaptation. To explore this possibility, we tested vibrotactile and ICMS detection thresholds at the end of a subset of the visits (**Fig. S7A**) and compared them to detection thresholds obtained at the beginning of the visit (~3 hours difference). We found no systematic difference in the pre- and post-session thresholds, confirming that the effects of the conditioning stimuli are not artifacts of within-visit changes in the participant’s sensitivity to vibration or ICMS. The median threshold shift for vibration was −0.5 dB (*p* > 0.05, **Fig. S7B**). Similarly, the median threshold shift for ICMS was 0.3 dB (*p* > 0.05, **Fig. S7B**).

Further, when considering only the magnitude of baseline detection threshold adaptation, sensitivity to vibration did not significantly change (p > 0.05) although sensitivity to ICMS did show a significant, but minor, change (median: 0.4 dB, *p* < 0.05, **Fig. S7C**).

To determine if baseline detection threshold shifts are correlated with shifts induced by non-target subthreshold stimulation, we computed the Pearson’s correlation coefficient and found no significant correlation between the conditions (*p* > 0.05, **Fig. S8**)

## DISCUSSION

By investigating the unexplored consequences of co-located touch perceptions in the hand stemming from different mechanisms of neural activation, we found that the simultaneous delivery of cortical stimulation and skin vibration can lead to changes in sensitivity to the tactile percepts elicited by either stimulus. Specifically, detection of skin vibration is enhanced in the presence of a subthreshold ICMS being delivered to a somatotopically overlapping region of the somatosensory cortex.

In accordance with previous work demonstrating that tactile sensitivity can be improved with non-invasive sensory noise signals through stochastic resonance (*21*, *22*), our results establish that a similar effect is achievable through the use of direct cortical stimulation. It is unclear if direct cortical activation manifests in altered neuron excitability (*23*) and thus increased tactile sensitivity in the periphery; however, it is clear that these findings indicate a potential role for ICMS in enhancing tactile sensitivity in individuals with sensory impairments.

Directly stimulating the somatosensory cortex was sufficient for decreasing vibrotactile stimulation detection thresholds, which could suggest that ascending information from peripheral inputs is not necessary to alter one’s perceptual sensitivity to touch. Alternatively, ICMS in the somatosensory cortex could recruit subcortical structures (e.g., cuneate or thalamus) that modulate tactile sensitivity. Regardless, the ability to reduce detection thresholds can be influenced by the direct activation of somatosensory cortex neurons.

### Effects of non-target subthreshold sensory stimuli on tactile perceptions

While subthreshold ICMS has the potential to significantly enhance sensitivity to tactile activity on the skin, a subthreshold vibration stimulation did not modulate detectability of ICMS (**Fig. 2A**). This conflicts with prior studies that observed increased excitability in motor axons after subthreshold peripheral stimulation (*24*, *25*). One possible reason for the lack of perceptual modulation during subthreshold skin vibration could be that the stimulus was not large enough to activate mechanoreceptors in the skin. Although we posit that the indentation from the tactor on the skin would lead to an increased mechanoreceptor response, it is unclear the degree to which a response to subthreshold stimulation manifests in the somatosensory cortex where ICMS was delivered.

### Effects of non-target suprathreshold sensory stimulation on tactile perceptions

We found that the simultaneous presence of a perceptible, suprathreshold ICMS did not significantly alter the detectability of the peripheral skin vibration (**Fig. 2A**); however, when measuring ICMS detection thresholds, the presence of a suprathreshold skin vibration *decreased* ICMS sensitivity (i.e., the detection thresholds increased) (**Fig. 2B**). It is unclear why this effect of decreased sensitivity to touch occurred only when ICMS was disrupted by suprathreshold skin vibration. One explanation could be that direct cortical stimulation to evoke touch sensations likely does not engage the same subcortical pathways as does touch on the skin, thus making perceptions from cortical stimulation more susceptible to modulation by distracting stimuli. In prior work, we found that when vibration and ICMS were approximately matched in intensity and were presented simultaneously, the participant occasionally reported only feeling vibration, suggesting a dominance of vibratory sensations over ICMS (*17*).

### Proximity of target and non-target stimuli is important

As the cortical distance between the maximally overlapping projected field of subthreshold ICMS and the vibration location increased – that is, as the ICMS electrodes were farther away from the projected field that overlapped with the skin vibration location – the magnitude of the enhanced tactile sensitivity decreased. The impact of subthreshold ICMS location on the altered vibration sensitivity is consistent with previous tactile masking studies that suggest that co-location of stimuli is necessary (*26*, *27*). In another study, providing low levels of ICMS to neighboring neural tissue was enough to increase detectability of a target ICMS signal (*13*). However, our results provide evidence for a gradual fall-off in tactile sensitivity enhancement as the neural stimulation gets farther away from the target location. The largest effect was observed when the ICMS signal was delivered to a neural region that perceptually overlapped with the skin vibration; as the ICMS location in the somatosensory cortex became farther away, the sensitivity enhancement decreased, leading to a negligible effect on sensitivity if more than 7 mm away (**Fig. 3B**, **Fig. 4B**). It is clear that the proximity of the non-target sensory stimulus to the target neural region is a driving factor in modulating sensitivity.

### Modulation of tactile sensitivity is not sustained

The modulation of tactile sensitivity during a sensory masking stimulus was immediate and did not persist over the course of an experimental visit (**Fig. S7**), suggesting that any altered performance during a tactile detection task is dependent on simultaneous delivery of the non-target stimulus. Prior work has demonstrated that adaptation to vibration (*28*) and ICMS (*29*) can occur if the stimuli are delivered for several seconds; however, here, the sensory stimuli were delivered well below the timescales generally observed to cause adaptation.

Although our results were repeatable over seven months and across multiple sites on both hands, exploring the effect of enhanced tactile sensitivity through cortical stimulation in additional participants is still needed. These results cannot explain the underlying mechanism of modulated tactile sensitivity during the simultaneous delivery of multiple tactile stimuli; however, it is clear that perceptual improvements in touch detection are enabled by cortical stimulation.

In conclusion, we showed that delivery of subthreshold ICMS to somatosensory cortex increased detectability of peripheral vibratory stimuli. This perceptual enhancement effect is dependent on ICMS delivery to overlapping or closely neighboring neural tissue that encodes touch in the region of the hand receiving vibration and degrades as the target and non-target stimuli get farther apart. The results of this study demonstrate the feasibility for enhancing tactile sensitivity of intact peripheral touch pathways degraded by disease or injury.

## MATERIALS AND METHODS

### Human participant

A male participant with a C5 motor / C6 sensory spinal cord injury participated in the study. The participant was implanted with six microelectrode arrays (Blackrock Neurotech; Salt Lake City, UT, USA) in sensorimotor regions of the brain (*16*, *19*). In the somatosensory cortex, there were two arrays in the left hemisphere and one array in the right hemisphere. Similarly, in the motor cortex, there were two arrays in the left hemisphere and one array in the right hemisphere (*16*, *19*). The participant had some retained motor functionality in the upper arms and wrist. Peripheral touch sensations were considered largely intact by the participant as well as by clinical assessments before implantation (*19*).

The study was part of a registered clinical trial (NCT03161067) and was conducted under an Investigational Device Exemption (IDE 170010) issued by the Food and Drug Administration (FDA). The study was reviewed and approved by the Johns Hopkins Institutional Review Board, the FDA, and the Naval Information Warfare Center (NIWC) Human Research Protection Office.

### Intracortical microstimulation

ICMS was delivered using biphasic, charge-balanced, cathodic-first pulses. The total width of each pulse was 500 μs (200 μs for each phase with a 100 μs interphase delay) and stimulation frequency was set to 100 Hz, which were values also used in previous studies (*16*, *17*, *30*, *31*). The pulse amplitude did not exceed 80 μA on any electrode. The projected fields for ICMS-evoked tactile percepts for each electrode were verbally mapped out: the participant described their locations using a hand map (*16*). In this study, ICMS was delivered to two electrodes at a time, each with similar projected fields, for each target site on the hand. Stimulation through two electrodes increased the perceived intensity of the tactile stimulus (*32*), making it easier for the participant to detect. ICMS was controlled using a custom MATLAB (MathWorks, Inc.; Natick, MA, USA) interface that communicated with a CereStim R96 (Blackrock Neurotech) to deliver ICMS pulses. The ICMS sensation was generally described as a pressure on the sites explored in this study, although the participant reported sensations of tingling as a result of ICMS through some of the other electrodes (*16*).

### Skin vibration

Skin vibration was delivered to each hand region using an electromechanical tactor (C-3, Engineering Acoustics, Inc.; Casselberry, FL, USA). The vibrotactile stimulus frequency was set to 300 Hz and the tactor was attached to the skin using medical tape. A custom MATLAB interface was developed to send sinusoidal signals through a computer’s audio port, which were amplified (Pyle, PTA4 Stereo Power Amplifier) to drive tactor indentation. Vibration intensity was controlled by varying the displacement of the tactor, which changed the indentation depth into the skin. The participant could not hear any noise coming from the tactor during the experiments.

### Estimating detection thresholds

Detection thresholds for skin vibration and ICMS were estimated using a two-alternative forced-choice (2AFC) paradigm. In this paradigm, the 0.5 s target stimulus was delivered in one of two intervals – each 0.5 s in duration with a 1 s delay between each interval – and the participant verbally reported in which interval they perceived the stimulus. Auditory cues were used to indicate the start of each interval, and the auditory tone frequency was different for the two intervals. When delivering a non-target stimulus, the non-target stimulus was delivered for 0.5 s in both intervals of the 2AFC task while the target stimulus was randomly delivered to only one of the two intervals (**Fig. 1D**).

We used the three down – one up (3D-1U) adaptive staircase procedure to estimate the detection threshold (*33*, *34*). We implemented the adaptive staircase method by decreasing the stimulation pulse amplitude until there was no detectable percept, at which point a ‘reversal’ occurred. The amplitude of the target stimulus was increased using a constant amplitude step size every time the participant did not correctly identify the 2AFC interval that contained the stimulus. We used a 5 µA amplitude step size for ICMS, which corresponded to approximately between 0.6 – 1.9 dB (median: 1.1 dB) when close to the detection threshold. For vibration, we used a step size that ranged from 0.76 mV to 51 mV, which corresponded to approximately between 0.5 – 2.0 dB (median: 1.1 dB) when close to the detection threshold. After the first reversal, the amplitude step size of the stimulus did not change for a given trial. Three correct responses (i.e., the participant correctly identified the 2AFC interval that contained the stimulus) in a row at a given stimulus resulted in a decrease in the target stimulus amplitude. A “reversal” occurred whenever the sign of the staircase slope changed. For each condition tested, the detection threshold was then calculated as the average stimulus amplitude at all the reversals (excluding the first reversal value). For each condition tested, we used six reversals to calculate the detection threshold. The 3D-1U adaptive staircase procedure with a 2AFC task estimates the stimulus amplitude for 79.4% detection probability (*33*).

When analyzing results from single sessions (**Fig. 1F** and **Fig. 3B**), a psychometric function was fit to the detection probabilities for each amplitude of the target stimulus on a given trial using the following logistic function:

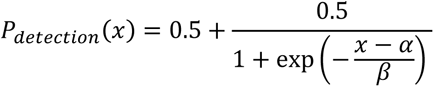

where *x* is the target stimulus amplitude, α is the halfway point of the psychometric curve (75% detection probability), and *β* is the steepness of the curve. Because the adaptive staircase procedure generally undersamples extremely high and low amplitudes that are very far away from the perceptual threshold, we assume that 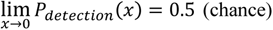 and 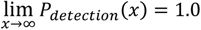, which is reflected in the psychometric function used to fit the data. The adaptive staircase procedure does not sample each stimulus amplitude level the same number of times, therefore when fitting the psychometric function, we weighted the data points based on the number of presentations at each amplitude level. As a result, stimulus amplitudes that were more heavily sampled (e.g., values close to the perceptual threshold) were given more weight when fitting the psychometric curve (e.g., **Fig. 1F**, **Fig. 3B**). Detection thresholds estimated from the psychometric curve were defined as the 79.4% probability in order to match the same level of probability of detection estimated by the adaptive staircase 2AFC procedure.

Thresholds and stimuli levels used during a testing session were expressed in dB using the following conversion:

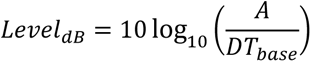

where *A* is the stimulus amplitude level and *DT_base_* is the baseline detection threshold measured for the stimulation modality at the target site at the beginning of that testing session. For context of scale, the just-noticeable difference (JND) of a sensory signal can be approximated as 1 dB (*35*), although smaller JND (~0.7 dB) for skin vibration have been reported (*36*).

### Bootstrapping experimental detection threshold data

From each adaptive staircase session, which produced a single detection threshold estimate, we simulated an additional 500 repetitions using a bootstrap method. The probabilities of detection were calculated for each stimulus level from the experimental psychophysical sessions trials and used to simulate the responses by implementing the same rules employed by the 3D-1U adaptive staircase approach, which produced an estimated detection threshold at the 79.4% probability level. For each experimental session, we then counted the proportion of simulated detection thresholds for that session that fell above or below the baseline value (**Fig. S1**). We used a significance level of 0.05 to determine if the experimentally measured detection threshold was significantly different from the baseline value. For example, if fewer than 5% of the bootstrapped detection thresholds for a given experimental trial were above the baseline value, then that experimentally measured detection threshold was considered to be significantly lower than baseline.

### Experimental procedures

For clarity, we use the term “visit” to describe a physical visit by the participant to the lab and “session” to describe a single psychometric sweep using the 2AFC adaptive staircase. The term “trial” is used to describe one presentation of one stimulation amplitude within a 2AFC adaptive staircase session. For each visit, the participant was seated with hands supinated and resting in his lap. Similar to our prior work, the vibrating motor was attached to his hand using medical tape, in a region that overlapped with an ICMS projected field (*16*, *17*, *30*). During each testing visit, we measured either ICMS or vibration detection thresholds (i.e., a “target stimulus”). The participant was told which stimulation modality was the target stimulus (i.e., ICMS or haptic vibration). ICMS was the non-target signal during vibration detection threshold experiments and vice versa. In prior work, we showed that the participant was able to identify the two stimuli (haptic vibration and ICMS) when delivered simultaneously with up to 93% accuracy (*17*).

We first measured the baseline detection threshold for the target stimulation modality and then repeated the psychophysical tests, using the adaptive staircase approach, to measure detection thresholds during different non-target tactile stimulation conditions. The non-target tactile stimulation conditions were 1) subthreshold (i.e., participant could not perceive the non-target stimuli), 2) suprathreshold with low amplitude (i.e., the participant verbally described that he could faintly feel the non-target stimuli), 3) suprathreshold with high amplitude (i.e., participant could easily feel the non-target stimuli), and 4) suprathreshold with a random amplitude (i.e., participant could feel the non-target stimuli but the intensity was randomized) (**Fig. 1E**).

In all cases, the non-target stimuli were the opposite modality of the target stimulus and for all conditions the non-target stimulus was delivered on each phase of the 2AFC task. In some experimental blocks, only one of the non-target stimulus amplitudes (either low or high) was presented on each phase of the 2AFC task; in other experimental blocks, the amplitude of the non-target stimulus varied randomly across stimulus intervals. The “suprathreshold with a random amplitude” condition was used to verify that the participant was not relying on the combined ICMS and haptic vibration intensity to make a decision. Results were combined across all suprathreshold conditions because the low and high amplitude non-target stimuli had statistically indistinguishable effects on detectability (**Fig. S4**).

To measure any adaptation effect of the baseline sensitivity after the combined modality sensory stimulation, we re-measured the baseline detection threshold for the target stimulation modality at the end of some sessions (n = 6 for vibration, n = 8 for ICMS).

Across all experiments, we measured a total of 117 unique detection thresholds across all sites and conditions over a period of seven months (**Table S1**).

### Measuring ICMS proximity

To measure the effect of subthreshold ICMS location on vibration detection thresholds, we performed additional experiments by stimulating electrodes in the cortex that did not elicit tactile percepts in the same region as vibration. For example, vibration thresholds were measured on the thumb while ICMS was applied to the ring finger. We tested 11 different sites across both hands.

We calculated the physical linear distance between electrode groups used to deliver the non-target ICMS signal by using the known size of each microelectrode array implant (4.2 mm x 2.4 mm) as well as the spacing between electrodes (420 μm). To calculate distance between neural regions covered by the same microelectrode array, the centroid of each electrode group, which was always two electrodes except for the left index finger with one electrode, used during ICMS activation of hand regions was calculated and used to approximate distance between the target neural region (e.g., thumb fingertip) and the non-overlapping neural region (e.g., ring fingertip) (**Fig. 4A**). For finding distances between electrode groups that were on different microelectrode arrays, we used intraoperative images of array placement in cortex to measure the linear distance between the approximate positions of the electrode group centroids. The distances ranged from 0.63 to 7.1 mm. It should be noted that this two-dimensional distance measurement does not incorporate differences in height between electrode groups. As in, the distance approximation assumed that each electrode group resided on the same plane.

### Measuring ICMS projected field size

Projected field size and location were verbally reported by the participant, who was given a hand map with predefined segments (*16*, *37*). Projected fields were mapped during delivery of ICMS to single electrodes using amplitudes ranging between 60 and 80 µA. The full mapping of projected fields for this participant are reported in (*16*). The size of the projected field for each electrode was calculated as the combined areas of all segments of the hand map in which a tactile sensation was reported during ICMS. Even though ICMS projected fields were recorded and observed to be consistent over a two year period (*16*) and percepts are capable of being elicited by the implants for at least four years (*38*), for this study we only used the ICMS projected fields collected during the study period. Because projected fields from ICMS can be assumed to be additive (*20*), we combined the projected fields from individual electrodes when delivering ICMS through multiple electrodes. The projected field size for each stimulation location was normalized against the largest projected field area that was reported in this study.

### Data analysis

All analyses and bootstrapping simulations were performed using MATLAB. A one-sample Wilcoxon signed-rank test was used to determine if the median between measured detection thresholds for a given condition and the baseline was nonzero (i.e., they differed from baseline detection threshold values). This test was chosen for comparing distributions to baseline values because we wanted to compare the detection thresholds measured in presence of non-target stimuli to their respective baseline detection threshold measurements taken at the beginning of each visit.

The Pearson correlation coefficient of detection threshold effects during subthreshold and suprathreshold non-target stimulation conditions was calculated with the MATLAB *corrcoef* function and by using data generated from within the same testing visit and same location on the participant’s hand. The statistical *p*-value of the correlation coefficient was determined from the correlation’s *t*-statistic with *n-2* degrees of freedom, where *n* was the number of detection threshold pairs.

A Wilcoxon rank-sum test was used for comparing distributions of detection thresholds collected across different conditions (e.g., subthreshold non-target ICMS versus suprathreshold non-target ICMS). This test was chosen for comparing two detection threshold distributions because we assumed the samples were independent across conditions. A binomial test was used to compare the number of detection thresholds that were significantly different than baseline to chance levels. Chance levels were calculated as the proportion of bootstrapped baseline detection threshold that were significantly different from the experimentally measured baseline detection threshold. Effect size was measured using *d_r_* (robust Cohen’s *d*) given its ability to appropriately estimate effect sizes from nonnormally distributed data (*39*, *40*). Unless noted, the violin plot whiskers represent the minimal and maximal values, the vertical lines indicate the first and third quartiles, the horizontal lines are means, and the white dots are the medians.

## Data Availability

All data produced in the present work are contained in the manuscript.

## Supplementary Materials

**Fig. S1:**
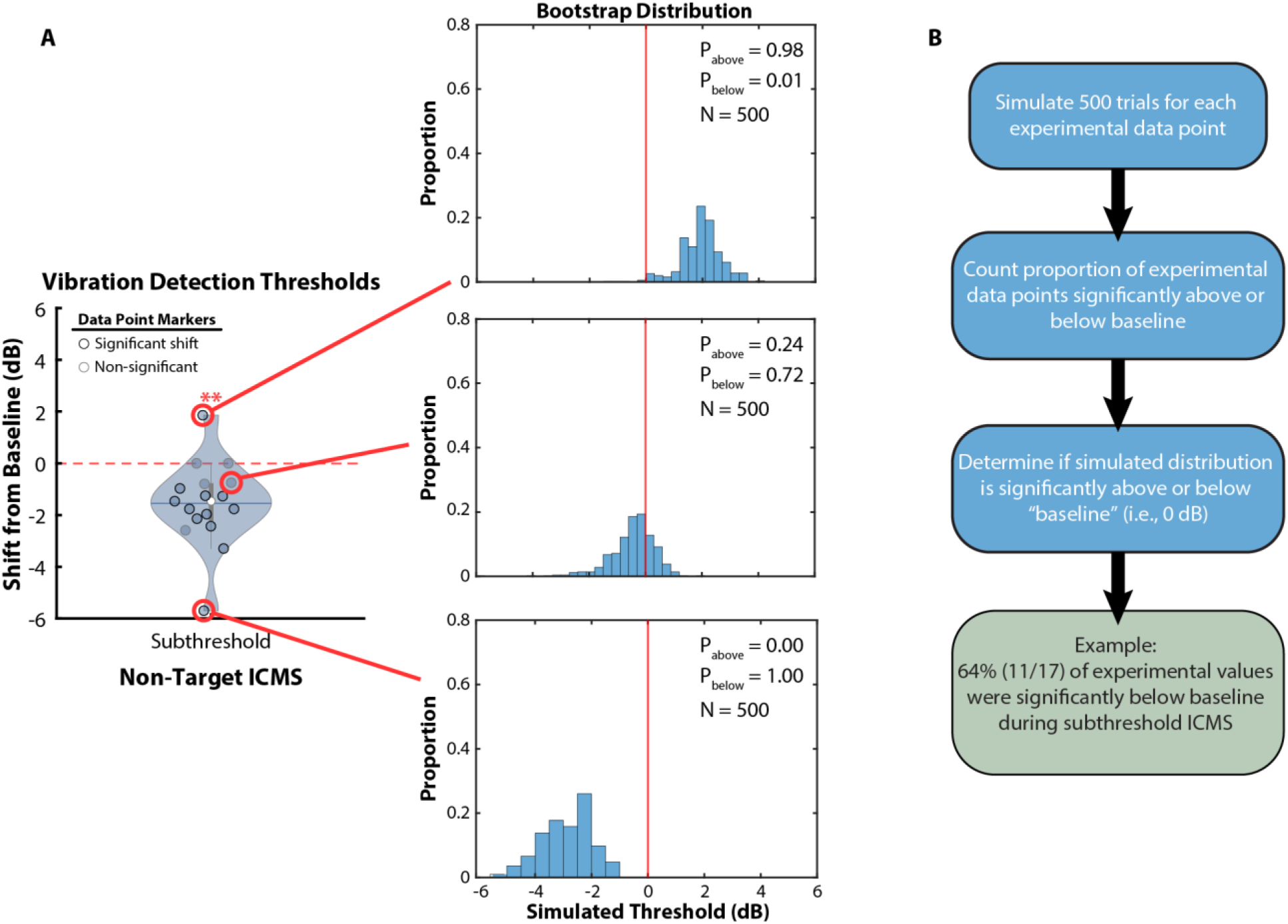
Bootstrap simulation to determine individual detection threshold significance. **(A)** A bootstrap analysis was performed by simulating 500 trials for each experimentally measured detection threshold. **(B)** The proportion of simulated detection thresholds above and below the baseline value were counted and used to determine the significance of each experimentally measured detection threshold. For example, if the simulated detection thresholds for a given experimental trial had less than 5% of the values above the baseline, that experimental data point was considered to be significantly less than the baseline detection threshold.

**Fig. S2:**
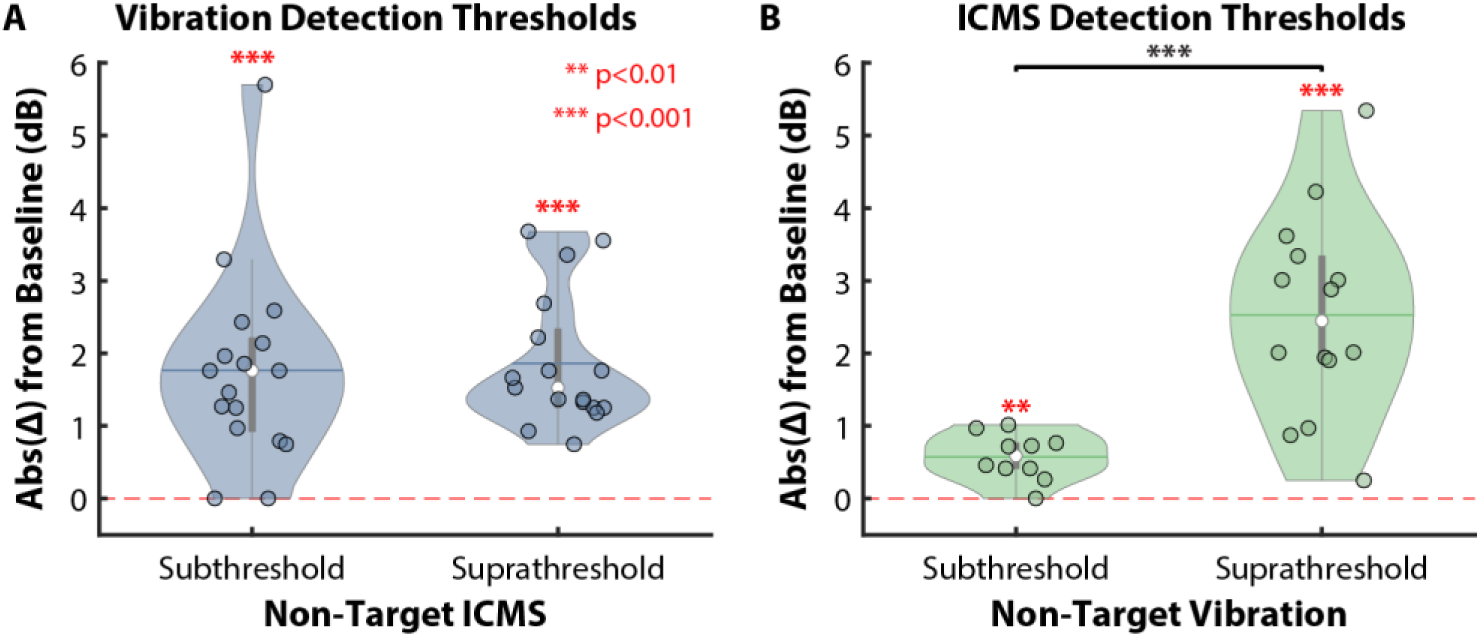
Non-target stimulation always shifted detection threshold. **(A)** When considering the magnitude of the change in skin vibration detection thresholds, there was a significant change compared to baseline during subthreshold (median: 1.8 dB, *p* < 0.001) and suprathreshold (median: 1.5 dB p < 0.001) ICMS; however, there was not a significant difference in the observed effect size between the two ICMS conditions (*p* > 0.05). **(B)** Similarly, simultaneous delivery of skin vibration caused an absolute change in ICMS detection thresholds during subthreshold (median: 0.6 dB, *p* < 0.01) and suprathreshold (median: 2.4 dB, *p* < 0.001) non-target vibration. There was an observed difference in effect size between the two non-target skin vibration amplitudes (*p* < 0.001).

**Fig. S3:**
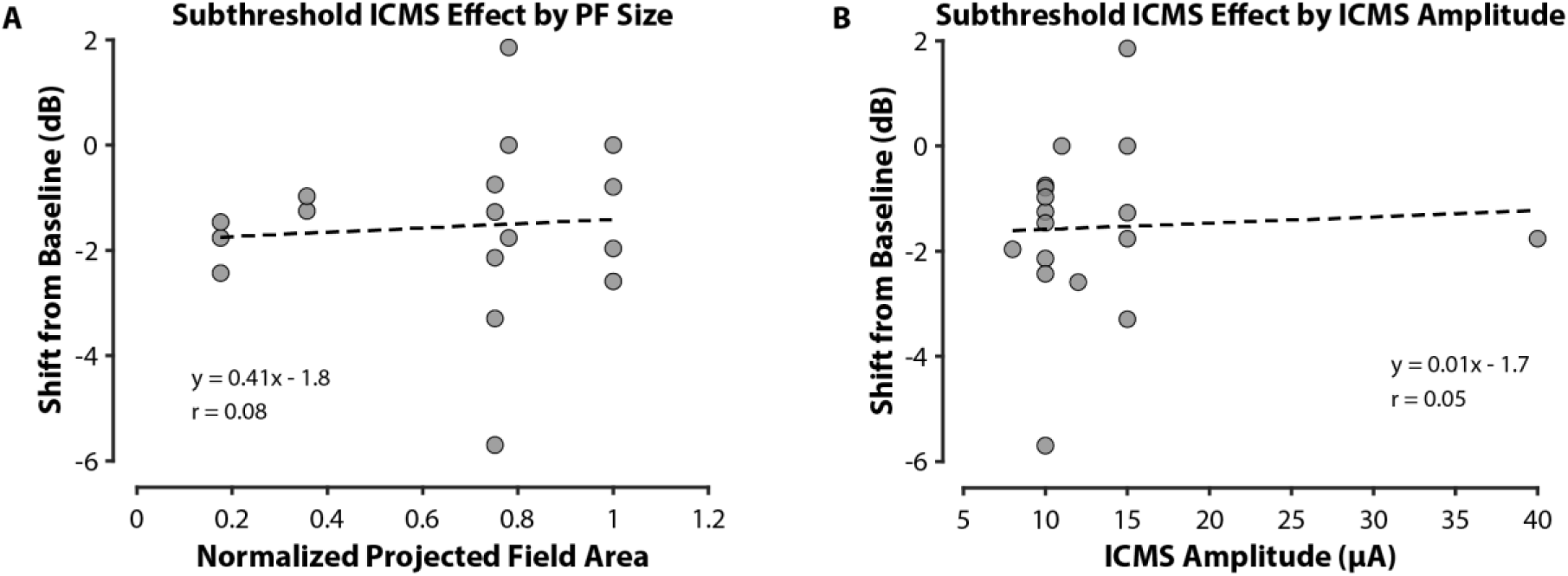
Subthreshold ICMS effect was not correlated with projected field size or ICMS amplitude. **(A)** The shift in vibration detection threshold from baseline during simultaneous delivery of subthreshold ICMS was not correlated with the normalized area of the projected field (PF) that corresponded to the neural region receiving ICMS (Pearson correlation coefficient *r* = 0.08, *p* > 0.05). **(B)** Similarly, vibration detection threshold shifts during subthreshold ICMS were not correlated with absolute ICMS amplitude (*r* = 0.05, *p* > 0.05)

**Fig. S4:**
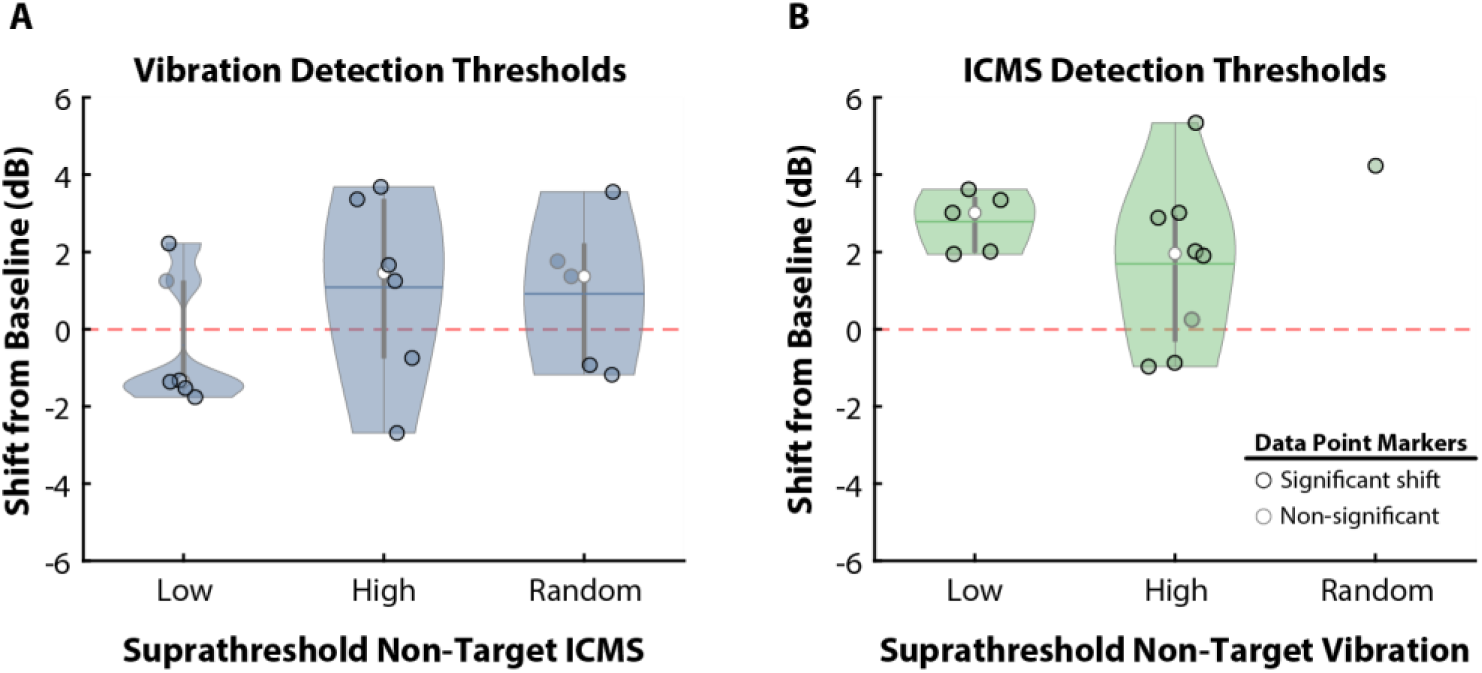
Detection thresholds did not vary significantly across different suprathreshold non-target stimuli conditions. The three different suprathreshold conditions were low (non-target stimuli were slightly perceptible), high (non-target stimuli were very perceptible), and random (non-target stimuli amplitudes were all perceptible but random on each phase of the 2AFC task). In the case of suprathreshold non-target stimuli, detection thresholds of target stimuli were similar across all three conditions (no significant difference in medians, *p* > 0.05) for both **(A)** skin vibration and **(B)** ICMS detection thresholds. **(A)** Skin vibration detection thresholds had median values of −1.3 dB (*p* = 0.56, *n* = 6), 1.5 dB (*p* = 0.31, *n* = 6), and 1.4 dB (*p* = 0.31, *n* = 5), compared to baseline, during low, high, and random non-target ICMS conditions and there was no significant difference between the conditions (p > 0.05). The number of trials significantly above baseline levels was 1/6 (16.7%), 4/6 (66.7%), and 1/5 (20%) for the low, high and random non-target ICMS conditions, respectively. The number of trials significantly below baseline levels was 4/6 (66.7%), 2/6 (33.3%), and 2/5 (40%) for the low, high, and random non-target ICMS conditions, respectively. **(B)** Median ICMS detection thresholds were higher than the baseline value for the low (3.0 dB, *p* = 0.063, *n* =5), high (2.0 dB, *p* = 0.078, *n* = 8), and random (4.2 dB, *n* = 1) suprathreshold non-target vibration conditions. The number of trials significantly above the baseline value was 5/5 (100%), 5/8 (62.5%), and 1/1 (100%) for the low, high, and random non-target vibration conditions, respectively. The number of trials significantly below the baseline value was 0/5 (0%), 2/8 (25%), and 0/1 (0%) for the low, high, and random non-target vibration conditions, respectively.

**Fig. S5:**
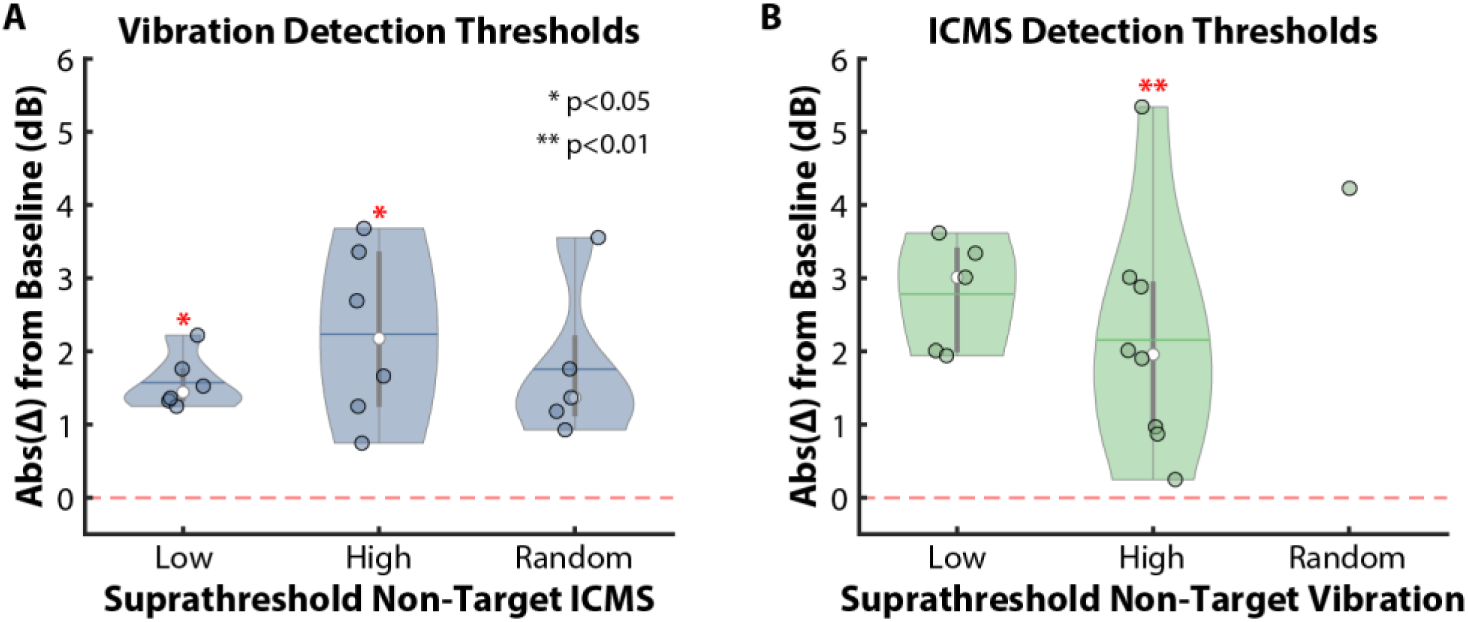
Magnitude of shift in detection threshold was not dependent on non-target stimulus amplitude. **(A)** The magnitude of the change in skin vibration detection thresholds was not significantly different (*p* > 0.05) when comparing the suprathreshold non-target ICMS conditions to each other; however, there was a significant shift from baseline in the Low (median: 1.4 dB, *p* < 0.05) and High (median: 2.2 dB, *p* < 0.05), conditions, but not the Random ICMS condition (median: 1.4 dB, *p* = 0.06). **(B)** There was not a significant difference (*p* > 0.05) in the magnitude of ICMS detection threshold changes when comparing the individual suprathreshold non-target vibration conditions to each other. The magnitude of the change in ICMS detection threshold was significant during the High non-target skin vibration condition (median: 2.0 dB, *p* < 0.01), but not in the Low (median: 3.0 dB, *p* = 0.06) or Random (median: 4.2 dB, *p* = 1.0) conditions.

**Fig. S6:**
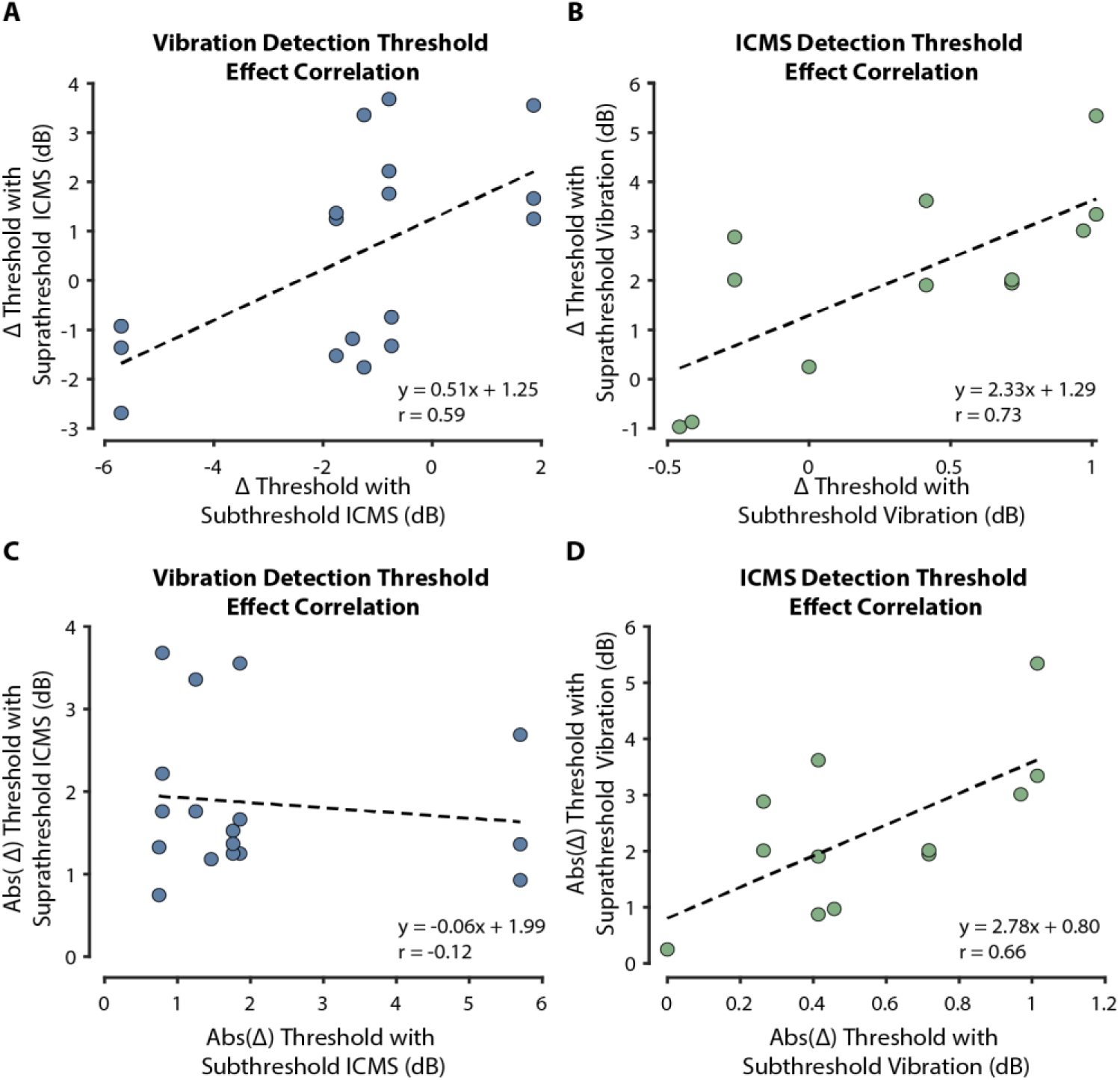
Subthreshold stimulation effect size was not correlated with suprathreshold stimulation effect size. Changes in **(A)** vibration (*r* = 0.59, *p* < 0.05) and **(B)** ICMS (*r* = 0.73, *p* < 0.01) detection thresholds were correlated between subthreshold and suprathreshold non-target stimulation conditions. When considering only the absolute change in detection thresholds, the size of the effect during suprathreshold non-target stimuli was not correlated with the effect size from subthreshold non-target stimuli in the case of **(C)** vibration detection (*r* = −0.12, *p* > 0.05); however, it was correlated for the **(D)** ICMS detection thresholds (*r* = 0.66, *p* < 0.05).

**Fig. S7:**
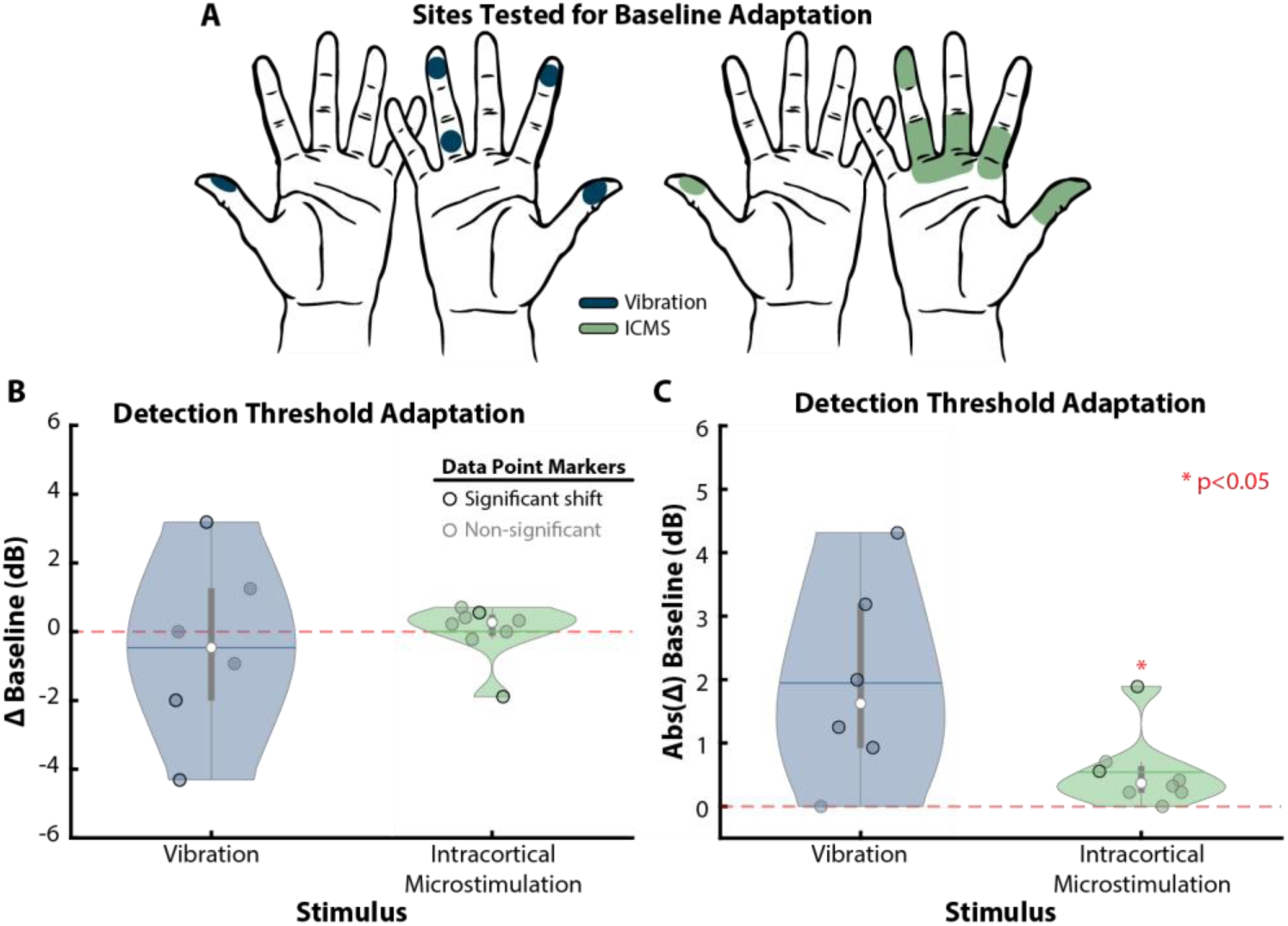
Baseline detection thresholds did not systematically shift over time. **(A)** Multiple sites were tested across both hands to determine if baseline detection of vibration or ICMS was altered within each visit. **(B)** At the end of each visit, detection thresholds were −0.5 dB (*p* = 0.81, *n* = 6, skin vibration) and 0.3 dB (*p* = 0.38, *n* = 8, ICMS) with respect to the thresholds measured at the start of the experimental visit. Detection thresholds without any non-target stimuli did not show systematic adaptation and were not significantly different from initial threshold estimates. **(C)** Vibration detection thresholds did not significantly change when considering only the magnitude of shift in baseline (median: 1.6 dB, *p* > 0.05); however, ICMS detection thresholds did have a small but statistically significant shift (median: 0.4 dB, *p* < 0.05).

**Fig. S8:**
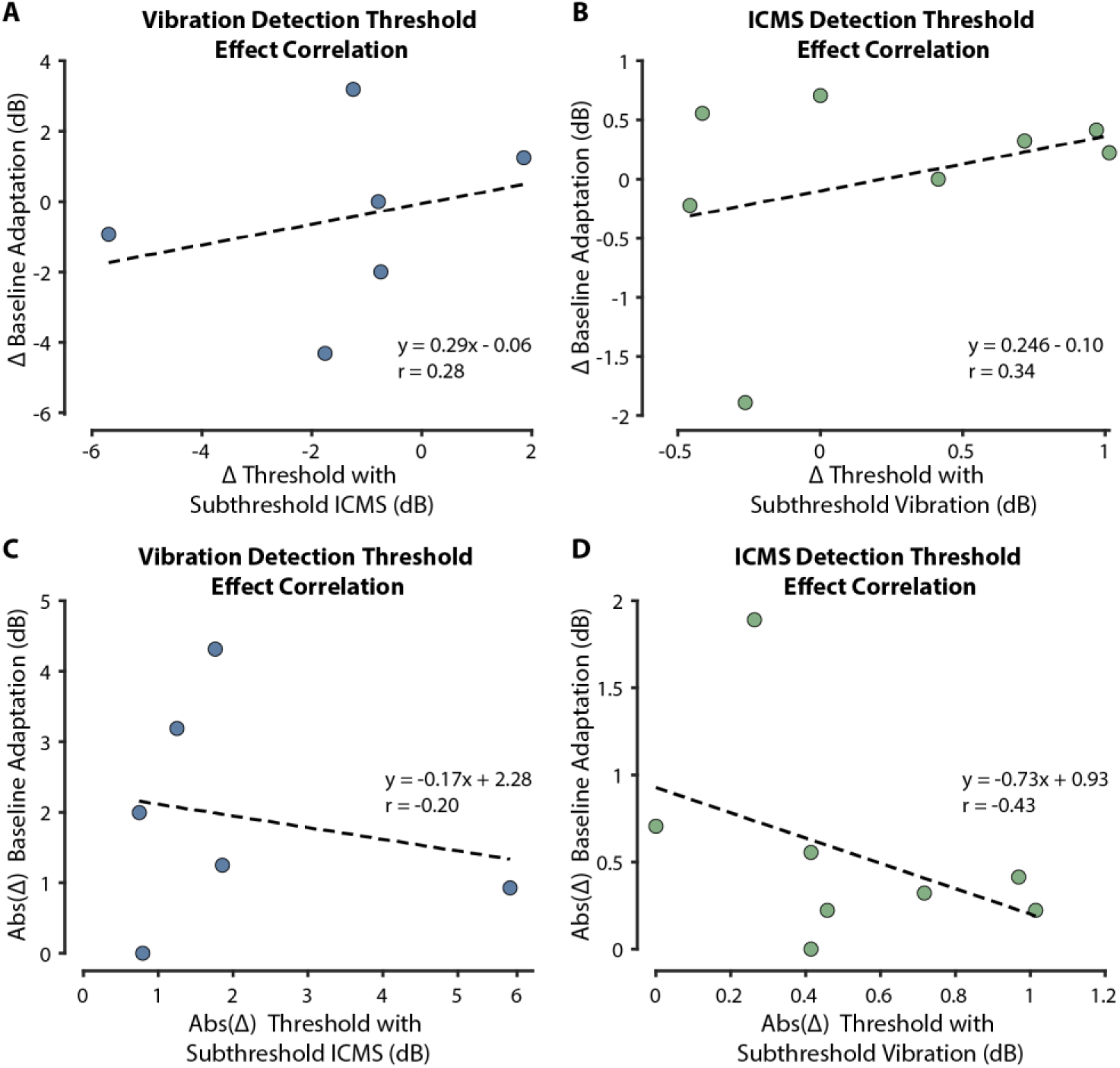
Size of subthreshold stimulation effect was not correlated with changes in baseline. The change in **(A)** vibration and **(B)** ICMS baseline detection threshold values was not correlated with the degree in which subthreshold non-target stimuli modulated detection thresholds. There was also no correlation when comparing the absolute change in **(C)** vibration and **(D)** ICMS detection thresholds with the absolute change in thresholds during the presence of subthreshold non-target stimuli. For all Pearson correlation coefficient, *r, p* > 0.05.

**Table S1:** Breakdown of sensory multiplexing testing conditions.

| **Non-Target**<br>**Stimulation Condition** | **Vibration**<br>**Thresholds (n)** | **ICMS**<br>**Thresholds (n)** |
| --- | --- | --- |
| None (Baseline) | 23 | 11 |
| None (Baseline Adaption) | 6 | 8 |
| Subthreshold | 17 | 10 |
| Offsite Subthreshold | 11 | 0 |
| Suprathreshold (low) | 6 | 5 |
| Suprathreshold (high) | 6 | 8 |
| Suprathreshold (random) | 5 | 1 |

## Author contributions

Conceptualization: BC, LEO, MSF, FVT, DPM

Methodology: BC, LEO, MSF, FVT, DPM, JMY, SJB

Investigation: LEO, BC, DPM, TMT, ASP, RWN, MSF, FVT

Analysis: LEO, BC, MFT, JMY

Visualization: LEO, BC, BAW

Funding acquisition: FVT, PAC

Project administration: FVT, MSF, PAC

Supervision: FVT, MSF, BAW, MAA, PAC

Writing – original draft: LEO, BC, SJB, MSF, FVT

Writing – review & editing: All authors

## Data and materials availability

The data needed to evaluate the claims made are included in the manuscript and supplementary materials. De-identified data is available upon request by qualified researchers through a materials transfer agreement.

## References

1. J. J. Collins, T. T. Imhoff, P. Grigg, Noise-enhanced tactile sensation. Nature 383, 770–770 (1996).

2. C. Wells, L. M. Ward, R. Chua, J. T. Inglis, Touch Noise Increases Vibrotactile Sensitivity in Old and Young. Psychol Sci 16, 313–320 (2005).

3. N. T. Dhruv, J. B. Niemi, J. D. Harry, L. A. Lipsitz, J. J. Collins, Enhancing tactile sensation in older adults with electrical noise stimulation. NeuroReport 13, 597–600 (2002).

4. A. Priplata, J. Niemi, M. Salen, J. Harry, L. A. Lipsitz, J. J. Collins, Noise-Enhanced Human Balance Control. Phys. Rev. Lett. 89, 238101 (2002).

5. P. Hänggi, Stochastic Resonance in Biology How Noise Can Enhance Detection of Weak Signals and Help Improve Biological Information Processing. ChemPhysChem 3, 285–290 (2002).

6. F. Moss, L. M. Ward, W. G. Sannita, Stochastic resonance and sensory information processing: a tutorial and review of application. Clinical Neurophysiology 115, 267–281 (2004).

7. F. Duan, F. Chapeau-Blondeau, D. Abbott, Weak signal detection: Condition for noise induced enhancement. Digital Signal Processing 23, 1585–1591 (2013).

8. L. R. Enders, P. Hur, M. J. Johnson, N. J. Seo, Remote vibrotactile noise improves light touch sensation in stroke survivors’ fingertips via stochastic resonance. J NeuroEngineering Rehabil 10, 105 (2013).

9. N. J. Seo, M. L. Kosmopoulos, L. R. Enders, P. Hur, Effect of Remote Sensory Noise on Hand Function Post Stroke. Frontiers in Human Neuroscience 8 (2014).

10. F.-G. Zeng, Q.-J. Fu, R. Morse, Human hearing enhanced by noise. Brain Research 869, 251–255 (2000).

11. K. S. Rufener, J. Kauk, P. Ruhnau, S. Repplinger, P. Heil, T. Zaehle, Inconsistent effects of stochastic resonance on human auditory processing. Sci Rep 10, 6419 (2020).

12. R. Kienitz, K. Kouroupaki, M. C. Schmid, Microstimulation of visual area V4 improves visual stimulus detection. Cell Reports 40, 111392 (2022).

13. L. E. Medina, M. A. Lebedev, J. E. O’Doherty, M. A. L. Nicolelis, Stochastic Facilitation of Artificial Tactile Sensation in Primates. J. Neurosci. 32, 14271–14275 (2012).

14. S. N. Flesher, J. E. Downey, J. M. Weiss, C. L. Hughes, A. J. Herrera, E. C. Tyler-Kabara, M. L. Boninger, J. L. Collinger, R. A. Gaunt, A brain-computer interface that evokes tactile sensations improves robotic arm control. Science 372, 831–836 (2021).

15. M. Armenta Salas, L. Bashford, S. Kellis, M. Jafari, H. Jo, D. Kramer, K. Shanfield, K. Pejsa, B. Lee, C. Y. Liu, R. A. Andersen, Proprioceptive and cutaneous sensations in humans elicited by intracortical microstimulation. eLife 7, e32904 (2018).

16. M. S. Fifer, D. P. McMullen, L. E. Osborn, T. M. Thomas, B. Christie, R. W. Nickl, D. N. Candrea, E. A. Pohlmeyer, M. C. Thompson, M. A. Anaya, W. Schellekens, N. F. Ramsey, S. J. Bensmaia, W. S. Anderson, B. A. Wester, N. E. Crone, P. A. Celnik, G. L. Cantarero, F. V. Tenore, Intracortical Somatosensory Stimulation to Elicit Fingertip Sensations in an Individual With Spinal Cord Injury. Neurology 98, e679–e687 (2022).

17. B. Christie, L. E. Osborn, D. P. McMullen, A. S. Pawar, T. M. Thomas, S. J. Bensmaia, P. A. Celnik, M. S. Fifer, F. V. Tenore, Perceived timing of cutaneous vibration and intracortical microstimulation of human somatosensory cortex. Brain Stimulation 15, 881–888 (2022).

18. G. A. Tabot, J. F. Dammann, J. A. Berg, F. V. Tenore, J. L. Boback, R. J. Vogelstein, S. J. Bensmaia, Restoring the sense of touch with a prosthetic hand through a brain interface. Proceedings of the National Academy of Sciences 110, 18279–18284 (2013).

19. D. P. McMullen, T. M. Thomas, M. S. Fifer, D. N. Candrea, F. V. Tenore, R. W. Nickl, E. A. Pohlmeyer, C. Coogan, L. E. Osborn, A. Schiavi, T. Wojtasiewicz, C. R. Gordon, A. B. Cohen, N. F. Ramsey, W. Schellekens, S. J. Bensmaia, G. L. Cantarero, P. A. Celnik, B. A. Wester, W. S. Anderson, N. E. Crone, Novel intraoperative online functional mapping of somatosensory finger representations for targeted stimulating electrode placement: technical note. Journal of Neurosurgery 135, 1493–1500 (2021).

20. C. M. Greenspon, N. D. Shelchkova, G. Valle, T. G. Hobbs, E. I. Berger-Wolf, B. C. Hutchison, E. Dogruoz, C. Verbarschott, T. Callier, A. R. Sobinov, E. V. Okorokova, P. M. Jordan, D. Prasad, Q. He, F. Liu, R. F. Kirsch, J. P. Miller, R. C. Lee, D. Satzer, J. Gonzalez-Martinez, P. C. Warnke, L. E. Miller, M. L. Boninger, A. B. Ajiboye, E. L. Graczyk, J. E. Downey, J. L. Collinger, N. G. Hatsopoulos, R. A. Gaunt, S. J. Bensmaia, Tessellation of artificial touch via microstimulation of human somatosensory cortex. bioRxiv, 2023.06.23.545425 (2023).

21. C. Well, L. M. Ward, R. Chua, J. T. Inglis, Touch noise increases vibrotactile sensitivity in old and young. Psychological Science 16, 313–320 (2005).

22. N. T. Dhruv, J. B. Niemi, J. D. Harry, L. A. Lipsitz, J. J. Collins, Enhancing tactile sensation in older adults with electrical noise stimulation. NeuroReport 13 (2002).

23. M. Bikson, M. Inoue, H. Akiyama, J. K. Deans, J. E. Fox, H. Miyakawa, J. G. R. Jefferys, Effects of uniform extracellular DC electric fields on excitability in rat hippocampal slices in vitro. The Journal of Physiology 557, 175–190 (2004).

24. H. Bostock, C. S.-Y. Lin, J. Howells, L. Trevillion, S. Jankelowitz, D. Burke, After-effects of near-threshold stimulation in single human motor axons. The Journal of Physiology 564, 931–940 (2005).

25. V. Deletis, J. Shils, J. Anso, E. Villar Ortega, L. Marchal-Crespo, K. A. Buetler, A. Raabe, K. Seidel, Effects of 10-kHz Subthreshold Stimulation on Human Peripheral Nerve Activation. Neuromodulation: Technology at the Neural Interface 26, 614–619 (2023).

26. R. T. Verrillo, G. A. Gescheider, B. G. Calman, C. L. Van Doren, Vibrotactile masking: Effects of one and two-site stimulation. Perception & Psychophysics 33, 379–387 (1983).

27. R. D. Gilson, Vibrotactile masking: Some spatial and temporal aspects. Perception & Psychophysics, doi: 10.3758/BF03209553 (1969).

28. M. Hollins, A. K. Goble, B. L. Whitsel, M. Tommerdahl, Time Course and Action Spectrum of Vibrotactile Adaptation. Somat Motor Res 7, 205–221 (1990).

29. C. L. Hughes, S. N. Flesher, R. A. Gaunt, Effects of stimulus pulse rate on somatosensory adaptation in the human cortex. *Brain Stimulation: Basic*, Translational, and Clinical Research in Neuromodulation 15 (2022).

30. L. E. Osborn, B. P. Christie, D. P. McMullen, R. W. Nickl, M. C. Thompson, A. Paware, T. M. Thomas, M. Anaya, N. E. Crone, S. J. Bensmaia, B. A. Wester, P. A. Celnik, G. L. Cantarero, F. V Tenore, M. S. Fifer, “Intracortical microstimulation of somatosensory cortex enables object identification through perceived sensations” in International Conference of the IEEE Engineering in Medicine and Biology Society (EMBC) (2021), pp. 6259–6262.

31. L. E. Osborn, D. P. McMullen, B. P. Christie, P. Kudela, T. M. Thomas, M. C. Thompson, R. W. Nickl, M. Anaya, S. Srihari, N. E. Crone, B. A. Wester, P. A. Celnik, G. L. Cantarero, F. V Tenore, M. S. Fifer, “Intracortical microstimulation of somatosensory cortex generates evoked responses in motor cortex” in International IEEE/EMBS Conference on Neural Engineering (NER) (2021), pp. 53–56.

32. J. T. Sombeck, L. E. Miller, Short reaction times in response to multi-electrode intracortical microstimulation may provide a basis for rapid movement-related feedback. J. Neural Eng. 17, 016013 (2019).

33. M. R. Leek, Adaptive procedures in psychophysical research. Perception and Psychophysics 63, 1279–1292 (2001).

34. F. Karmali, S. E. Chaudhuri, Y. Yi, D. M. Merfeld, Determining thresholds using adaptive procedures and psychometric fits: evaluating efficiency using theory, simulations, and human experiments. Experimental Brain Research 234, 773–789 (2016).

35. A. W. Mills, Lateralization of High-Frequency Tones. Citation: The Journal of the Acoustical Society of America 32, 132 (1960).

36. H. Pongrac, H. Pongrac, Vibrotactile perception: examining the coding of vibrations and the just noticeable difference under various conditions. Multimedia Systems 2007 13:4 13, 297–307 (2007).

37. S. N. Flesher, J. L. Collinger, S. T. Foldes, J. M. Weiss, J. E. Downey, E. C. Tyler-Kabara, S. J. Bensmaia, A. B. Schwartz, M. L. Boninger, R. A. Gaunt, Intracortical microstimulation of human somatosensory cortex. Science Translational Medicine 8, 361ra141 (2016).

38. C. L. Hughes, S. N. Flesher, J. M. Weiss, J. E. Downey, M. Boninger, J. L. Collinger, R. A. Gaunt, Neural stimulation and recording performance in human sensorimotor cortex over 1500 days. J. Neural Eng. 18, 045012 (2021).

39. J. Algina, H. J. Keselman, R. D. Penfield, An alternative to Cohen’s standardized mean difference effect size: A robust parameter and confidence interval in the two independent groups case. Psychological Methods 10, 317–328 (2005).

40. J. C.-H. Li, Effect size measures in a two-independent-samples case with nonnormal and nonhomogeneous data. Behav Res 48, 1560–1574 (2016).

